# Comorbidities associated with chronic kidney disease among young people living with HIV in Uganda. A nested case control study

**DOI:** 10.1101/2024.11.14.24317307

**Authors:** Esther M Nasuuna, Laurie A Tomlinson, Robert Kalyesubula, Barbara Castelnuovo, Nicholus Nanyeenya, Chido Dziva Chikwari, Helen A Weiss

## Abstract

**Introduction:** Chronic kidney disease (CKD) is often complicated by disorders in multiple body systems, associated with higher mortality and morbidity. Young people living with HIV (YPLHIV) have an increased risk of multisystem chronic comorbidities. However, there are few data describing comorbidities associated with CKD among YPLHIV.

**Methods:** We conducted a case-control study in seven ART clinics in Kampala, Uganda. Cases were YPLHIV (aged 10-24 years) diagnosed with CKD and controls were those without CKD. We collected data on demographic and clinical factors: blood pressure, fasting glucose levels, anaemia, electrolytes, parathyroid hormone, and cognitive impairment. We summarized the demographic and clinical factors and used logistic regression to estimate odds ratios (OR) and 95% confidence intervals for associations between CKD comorbidities, adjusted for age, sex and viral suppression.

**Results:** A total of 292 participants (96 cases and 196 controls) were recruited. Cases were mostly male (59.4% vs 36.5%), and younger (88.5% vs 46.4% aged <17 years) compared to controls. CKD was associated with having a detectable HIV viral load (OR=3.73; 95% CI 1.53-9.12) and proteinuria (aOR=4.19; 95% CI 2.28-7.72). CKD was also associated with low haematocrit, hypochloraemia, hyperphosphatemia, and high mean corpuscular volume. There was no evidence of an association of CKD with hypertension, anaemia, or stunting.

**Conclusion:** The pattern of comorbidities among YPLHIV with CKD is uncertain and difficulties may relate to difficulty determining true kidney function and normal ranges in this population. Further studies are needed to discern the pattern of CKD complications to improve management efforts and clinical outcomes.

## Introduction

Chronic kidney disease (CKD) is often complicated by disorders in multiple body systems as the kidneys play a pivotal role in homeostasis maintenance in the body [1]. With extensive damage and progressive decline in function, the kidney is unable to perform its many functions and other body systems are affected [2]. The most commonly affected systems are the haematological, cardiovascular, endocrine, and musculoskeletal [3, 4]. CKD comorbidities and complications increase with reducing kidney function and are associated with higher mortality, morbidity, and reduced quality of life [5].

In people with CKD, cardiovascular complications such as congestive cardiac failure, arrhythmias, and coronary artery disease are common, and are leading cause of mortality [6, 7]. Hypertension also commonly complicates CKD and leads to faster deterioration in glomerular filtration rate [8].

Anaemia is the commonest haematological complication of CKD resulting partially from the reduced production of erythropoietin in the damaged kidneys [4]. In the musculoskeletal system, disordered bone mineral metabolism of calcium, phosphorus and parathyroid hormone leads to osteodystrophy and osteoporosis that predispose to fractures and stunting in children [9-11]. The most common neurological complications are cognitive impairment, stroke, encephalopathy, autonomic and peripheral neuropathies [3, 12]. If these complications are undiagnosed and untreated, they will eventually lead to death [2].

Young people living with HIV (YPLHIV) have an increased risk of multisystem chronic comorbidities as a result of the HIV infection itself [13]. When YPLHIV develop CKD, the risk of comorbidities is increased due to the additional burden of gradual deterioration in kidney function [5, 14]. There is a poorly understood complex interplay between HIV and CKD that makes it difficult to determine if HIV or CKD or their combination is driving the development of the complications [15]. Current literature that is derived mostly from high income countries, describes the presentation and comorbidities of CKD in adults that are mostly caused by living with hypertension and diabetes, or in children with mostly congenital causes [16]. These comorbidities may differ from those in young adults living with HIV and CKD in LMICs, and we need to understand such differences, especially if they have clinical implications.

Kidney disease improving global outcomes (KDIGO) recommends the consistent documentation of all laboratory abnormalities in different populations with CKD to inform management practices and improve associated quality of life, morbidity and mortality [5]. Therefore, we sought to describe the comorbidities and complications and associations with kidney function in YPLHIV in Uganda.

## Materials and Methods

### Study design, setting and population

This case-control study with 96 cases and 196 controls was conducted in seven urban ART clinics in Kampala, Uganda between November 2023 and January 2024.

### Sampling

The cases were the YPLHIV (aged 10 to 24 years) diagnosed with CKD. This was defined as an eGFR below 90ml/min/1.73m^2^ on two separate occasions three or more months apart using cystatin C based Schwartz equation [17] without restriction for age. We elected to use this approach as sudden switch to adult formulae at age 18 have been shown to overestimate GFR and it has been recommended to use paediatric formulae for adolescents and young adults up to age 30 when concurrence between eGFR from adult and paediatric formulae happens [18, 19]. We used a cut off of 90ml/min/1.73m^2^ and not the standard <60ml/min/1.73m^2^ as we were studying a young population where GFR well above 60ml/min/1.73m^2^ would have been anticipated in health, and stage 2 CKD is diagnosed between 89 and 60ml/min/1.73m^2^ if there is evidence of persisting kidney damage [20-22]. Separately, we also described parameters among individuals with eGFR<60ml/min/1.73m^2^. Serum cystatin C was measured in a batch by particle-enhanced immunoturbidimetric assay on Roche Cobas C311 platform with Tina-quant Cystatin C Gen.2 [23].

The controls were the YPLHIV attending the same clinics with an eGFR equal to or above 90ml/min/1.73m^2^ determined with the same methods as the cases. We recruited two controls for each case.

### Study procedures

Participants were invited to the facility where they were interviewed about current medication and hospitalisation. Data were collected on the demographic variables (age, sex, religion, school status, tribe, address). School status was either in school or out if school, address was collected as either living in Kampala or out of Kampala. Clinical variables collected were: stunting according to the WHO reference standard, body mass index calculated as weight over height squared, mid upper arm circumference (MUAC) was measured using the UNICEF tapes according to the age of the participant, HIV viral suppression was defined as an HIV viral load below 1000 cells/ml, proteinuria was defined as either trace of 1+ and above on dipstick, albumin creatinine ratio (ACR) was measured in the laboratory and whether or not the participant was on a tenofovir disoproxil fumarate (TDF) based regimen.

The comorbidities assessed included hypertension, anaemia, impaired blood glucose, bone mineral disease, electrolyte imbalance, stunting and cognitive impairment. Blood pressure (BP) was measured digitally, using a paediatric cuff for the younger children. Participants were asked to sit down and relax, and their BP was taken on the left arm with an OMRON Upper ARM Blood Pressure Monitor HEM 7121J, (OMRON Healthcare Company Limited, Kyoto, Japan). The BP was taken twice, five minutes apart and the mean of the two readings taken as the final BP. They then underwent a cognitive function assessment using the Youth International HIV Dementia Scale (y-IHDS) [24].

Fasting blood glucose was measured using On Call Plus Blood Glucose Monitoring System (ACON Laboratories, Inc, San Diego, United States of America) and a venous blood sample was taken. The blood sample was analysed to determine values of serum electrolytes (potassium, chloride, bicarbonate, phosphate, calcium), kidney function (creatinine, blood urea nitrogen) measured on the Cobas 501/502 (Hitachi High technologies Corporation Tokyo, Japan). Haematological tests (complete blood count and serum ferritin) and parathyroid hormone were measured on the Cobas e601 (Hitachi High technologies Corporation Tokyo, Japan). All the samples were analysed in the College of American Pathologists (CAP) accredited MRC/UVRI laboratory in Uganda.

### Data analysis

Data were collected in REDCap (2019, Vanderbilt University, USA) and exported to STATA statistical software version 18 (STATA Corp USA) for analysis. To take account of the changing reference values across the age group of this study, comorbidities were defined as in Table 1.

**Table 1.** Reference Ranges for the assessed comorbidities among the participants.

| Variable | Reference range |
| --- | --- |
| Blood pressure (mmHg) | Elevated blood pressure was between 90<sup>th</sup> and 95<sup>th</sup> percentile for age, sex, and height.<br>Hypertension was above 95<sup>th</sup> percentile for children aged less than 13 years. Those above 13 years of age were considered hypertensive if the blood pressure was above 130/80 [8] |
| Fasting blood glucose (mg/dl) | Normal 70-100,<br>Prediabetes 100-125<br>Diabetic >126 [25]. |
| Haemoglobin (g/dl) | 11.5 <11 years ,<br>11.9 11 and 15 years, females >15 years<br>12.9 for males >15 years [26]. |
| Ferritin (ng/mL) | 12-300 for males >15 years<br>10-150 for females > 15 years<br>7-142 males and females 10-14 years (20). |
| Red blood cells | 4.7-6.1 for males<br>4.2-5.4 for females |
| Haematocrit | 0.35-0.48 for all ages [27] |
| Mean Corpuscular Volume (MCV) (fL) | 77.8-91.1 males 6-11 years<br>79.9-93.0 males 12-14 years<br>82.5-98.0 males 15-17 years<br>78.2-97.9 males >18 years<br>77.8-91.1 females: 6-11 years<br>79.9-93.0 females 12-14 years<br>82.5-98.0 females 15-17 years<br>78.2-97.9 females >18 years [28]. |
| Mean Corpuscular Haemoglobin (MCH) | 25-33 pg/cell for all ages and sexes [29]. |
| White blood cells (μL) | 4-10.5 X 10<sup>3</sup>/ for all ages and sexes |
| Platelets | 150-400 for all ages [27] |
| Parathyroid hormone (pg/ml) | 15-59 for 10 years,<br>15-68 for 11 to 17 years<br>15-65 for 8 to 24 years [30]. |
| Serum calcium (mg/dl) | 9.4-10.3 for 10 to 12 years<br>9.1-10.2 for males aged 12 to 24 years.<br>8.8-10 for females aged 12 and above [31]. |
| Serum Phosphate (mg/dl) | 3.6-5.8 for 10 -12 years,<br>2.3-4.5 for 12 -20 years [32].<br>2.5 to 4.5 for 21 to 24 years [33]. |
| Blood urea nitrogen (mg/dl) | 6-20 for 10-to-17-years<br>8-24 for males 18 to 24 years,<br>7-2 for females 18-24 years [34]. |
| Serum Potassium (mmol/l) | 3.6-5.2 for all ages [34]. |
| Serum chloride (mmol/l) | 102-112 for 10 to 17 years<br>98 to 107 for 18 to 24 years [34]. |
| Serum bicarbonate (mmol/l) | 21-29 in males<br>22-29 in females [34]. |
| Cognitive impairment | <10 on the (y-IHDS) [24]. |

### Statistical methods

Demographic and clinical characteristics were summarised by case/control status as number and percentage (categorical variables), and means and standard deviations, or medians and interquartile ranges (continuous variables) for the parametric data. A subgroup analysis of the cases that had eGFR <60ml/min/1.73m2 was also conducted. Associations of case/control status with comorbidities were assessed using logistic regression to estimate odds ratios and 95% CI. In modelling adjusted odds ratios for the association between case/control status and haematological or biochemical covariates we adjusted for age as a continuous variable, sex and viral suppression. P-values were estimated using likelihood-ratio tests. Post-hoc, to explore the extent to which differences in biochemical and haematological parameters were affected by adherence to anti-retroviral medication, we also conducted an analysis restricting to cases and controls who were virally suppressed. We also assessed the relationship between eGFR and selected haematological and biochemical factors using scatter plots and correlation coefficients to determine how they changed with changes in eGFR.

## Results

We recruited 96 cases and 196 controls. Cases were more likely to be male (59.4% vs 36.2%) and to be younger (88.5% vs 46.4%) with a lower mean age compared to controls (15.3 years vs 17.8 years). After adjusting for age and sex, cases were more likely to be virally non suppressed, have proteinuria, and to have ACR above 30mg/g (Table 2).

**Table 2:**
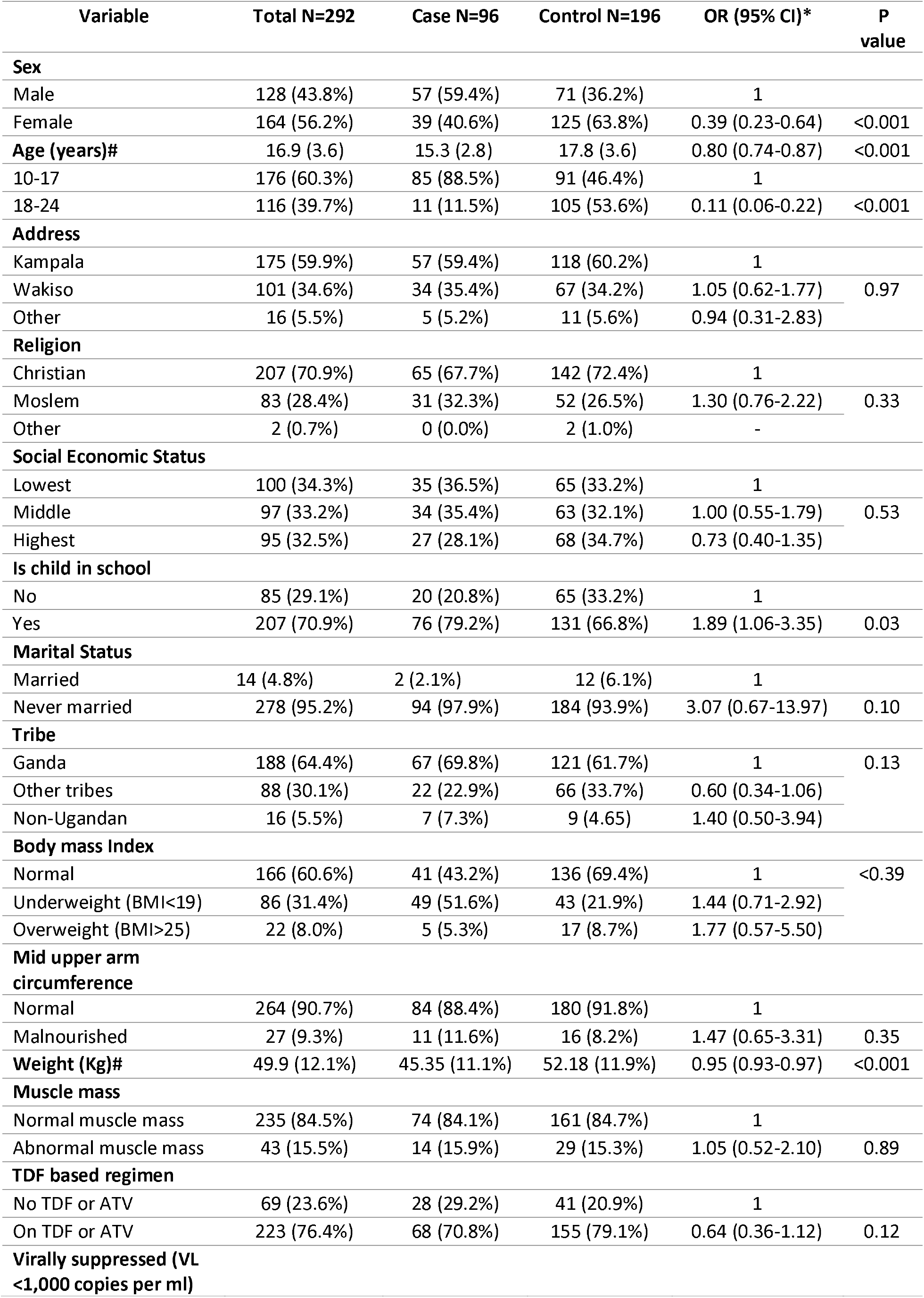
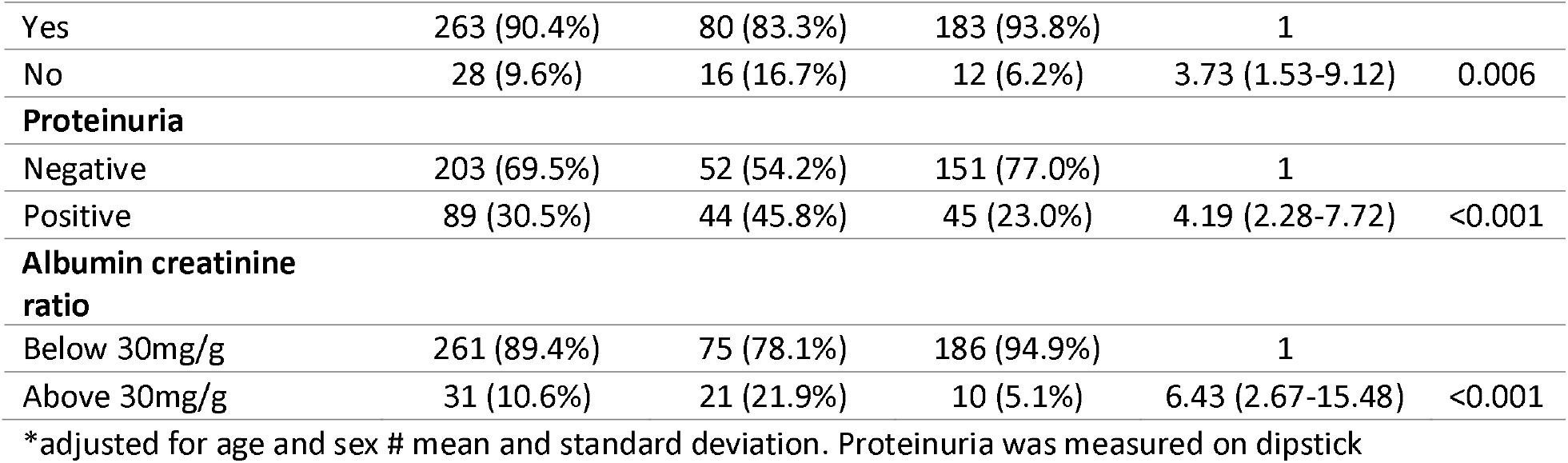
Distribution and associations of CKD with demographic and clinical characteristics among YPLHIV.

### Associations between CKD and comorbidities among YPLHIV

Cases had lower odds of having a high MCV (aOR=0.24, 95% CI 0.09-0.64), and higher odds of hyperphosphatemia (aOR=3.66, 95% CI 1.21 -5.84), and higher odds of hyperchloremia (aOR=2.55, 95%CI 1.36 -4.77), after adjustment for age, sex, and viral suppression. There was no evidence of an association between CKD and blood pressure, impaired blood glucose, other biochemical and haematological parameters and cognitive function. These results did not change when analysis was restricted to those who were virally suppressed (Table 3). Analysis of eGFR and selected variables showed no correlation between eGFR and those variables. (Figure 1). Of note, two participants had potassium levels below 3.5mmol/l and six had elevated levels above 6 mmol/l and 125 (42.8%) were acidotic. Results for the cases that had eGFR <60ml/min/1.73m2 are reported in supplementary tables 1 and 2.

**Table 3:**
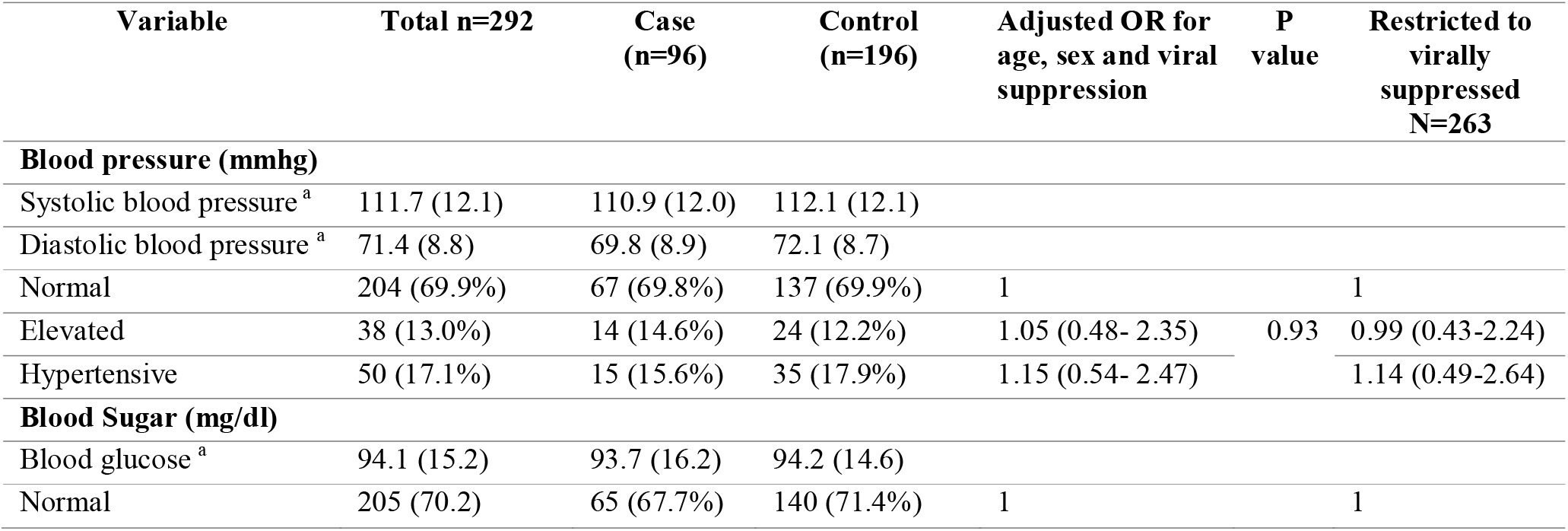
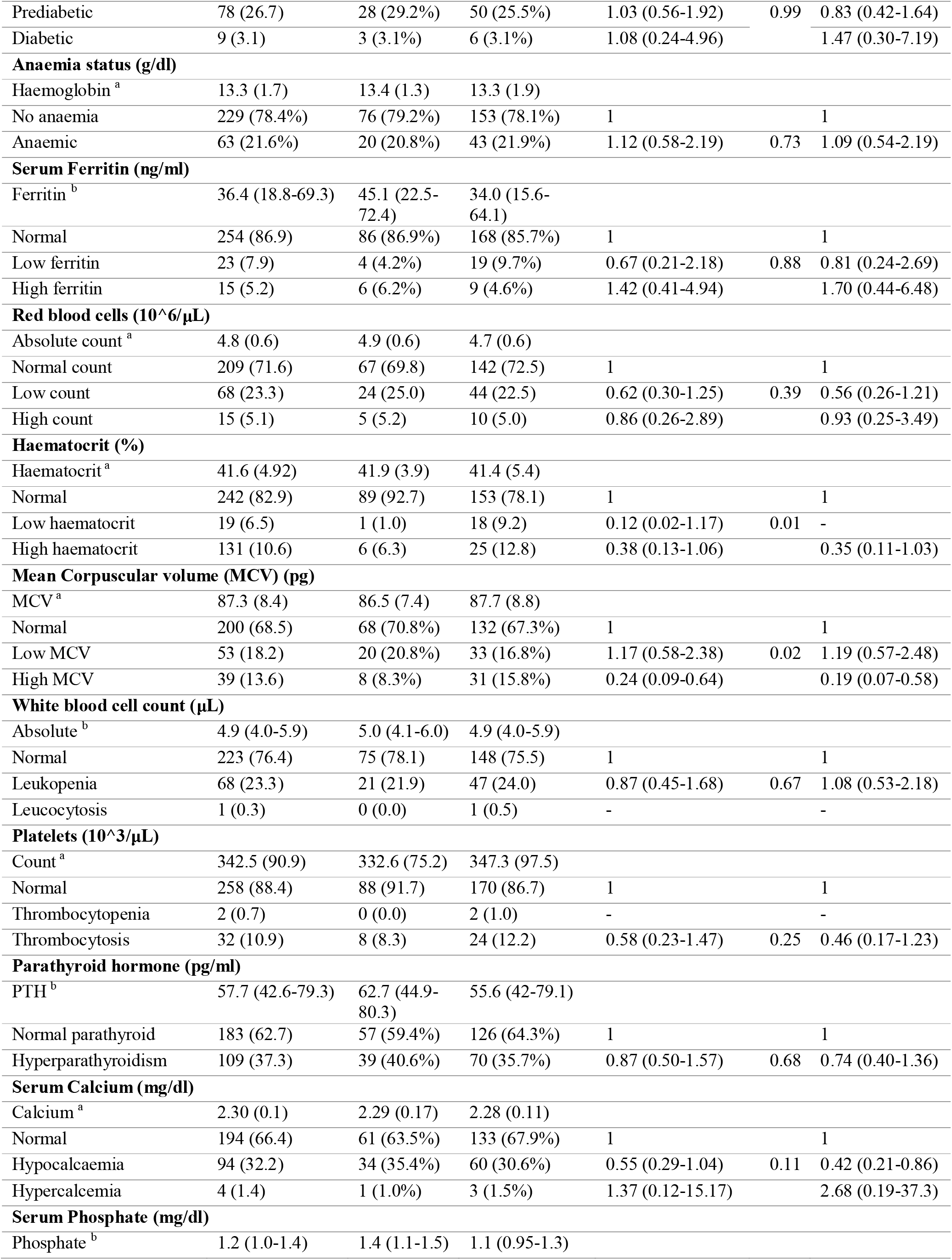
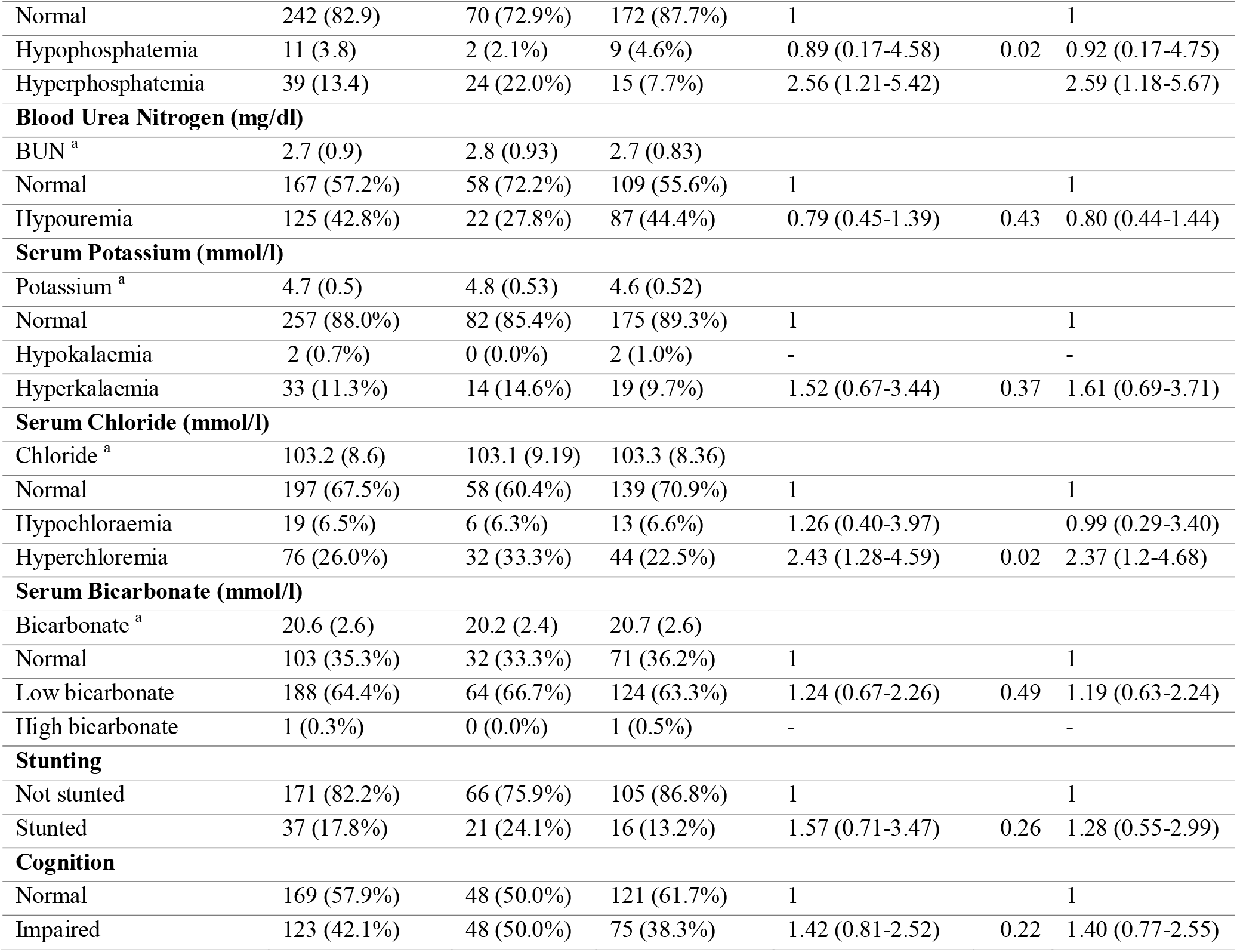
Distribution and associations of CKD with comorbidities among PLWH.

**Figure 1.**
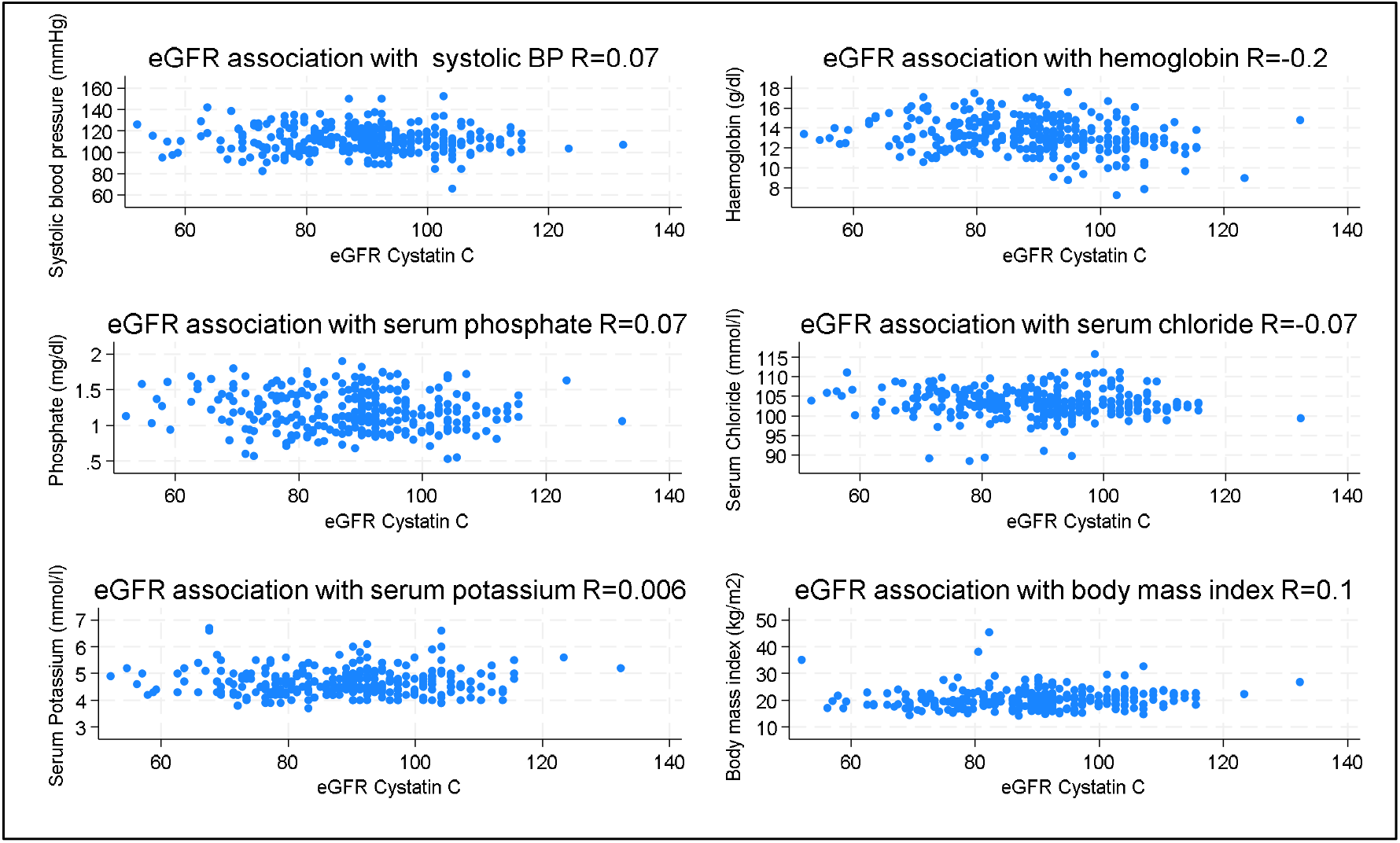
Relationship between eGFR and selected biochemical markers

### Cases with eGFR<60mls/min/1.73m^2^

Of the seven participants with eGFR <60ml/min/1.73m^2^, five were male, below 18 years of age and on a TDF based regimen, three had proteinuria and five had an ACR above 30mg/g. The mean systolic blood pressure was lower than that of the participants with eGFR below 90ml/min/1.73m^2^ and only one was hypertensive, two had prediabetic range blood glucose levels, one had anaemia, two had leukopenia, one had thrombocytosis and four had increased levels of parathyroid hormone and 5 had impaired cognition (Table 3).

## Discussion

We set out to assess comorbidities associated with CKD among YPLHIV in Uganda. YPLHIV with CKD were more likely than controls to have hyperphosphatemia and hyperchloremia and were less likely to have high MCV and high haematocrit. Although the prevalence of common comorbidities such as hypertension, anaemia, impaired glucose metabolism, cognitive impairment and stunting was high, they were not associated with having CKD among the YPLHIV in this study.

We included a sufficiently large number of cases and controls and assessed multi body systems for comorbidities. There is little literature on the comorbidities faced by young people with the double burden of CKD and HIV in SSA. This is surprising given that 90% of young people living with HIV reside in SSA and they are at high risk of developing complications of HIV such as CKD [35, 36].

Findings from the current study are thus discussed here in comparison with those in either young people living with CKD and not HIV, or with YPLHIV or adults with CKD in the general population.

We might not have observed substantial differences between YPLHIV with CKD and those without for a variety of reasons. 1) HIV itself causes similar complications to CKD in the cardiovascular, musculoskeletal, and haematological systems, including stunting [13]. The cases were more likely to have HIV viral non-suppression, an important difference since uncontrolled HIV can lead to more complications. 2) The cases (except for the seven with eGFR<60ml/ml/1.73m^2^) were still in the early stages of CKD with an eGFR between 60 and 90 ml/min/1.73m^2^, most complications develop with later stages of CKD [37]. 3) CKD status of the controls was determined only at baseline, any of them could have developed an eGFR less than 90ml/min/1.73m^2^ during the follow up time, making them a case and obscuring the differences. 4) Discerning participants with derangements in the biochemical and haematological markers was also challenging. Reference values for most of these markers were derived from American populations and those of African populations are mostly unknown [38]. Using these values could have led to misclassification of the YPLHIV and obscured associations [39].

This study shows that the cases have started developing complications and could get worsening kidney function and eventual kidney failure. The cases had hyperphosphatemia, but not low calcium. This was also true for the CKiD study in the United States of America that enrolled children with CKD, mostly of congenital origin. As GFR decreased, hyperphosphatemia and increased levels of PTH and eventually low levels of calcium emerged [40]. The cases in this study may be on their way to developing bone mineral disease which is associated with cardiovascular events, cognitive impairment and stunting [41]. The cases in our study had higher levels of serum chloride than controls. Serum chloride has been suggested as predictor of CKD progression and could show that the YPLHIV are in danger of CKD progression [42]. Of concern, we have identified people with both dangerously high and low potassium levels. We cannot exclude that these are due to haemolysis or analytical errors,) but this highlights the importance of better monitoring and clinical management of biochemical abnormalities in this vulnerable patient group.

YPLHIV with CKD have been shown to have atypical presentation patterns which are different from adults living with HIV where we draw most of the comparisons [13]. For example, common complications such as hypertension and anaemia were not associated with CKD in this study. A study in Nigeria on 24 children 6 to 17 years of age with CKD and age and sex matched controls found that prevalence of hypertension increased as the CKD stage increased [43]. The participants in this study were not living with HIV and had more advanced stages of CKD compared to our participants.

Haemoglobin was not associated with eGFR in this study. Anaemia is more prevalent in advanced stages of CKD with GFR less than 60 ml/min and since the YPLHIV were still in lower stages, this could explain the lack of association [44, 45].

There is still a lot that is unknown about the comorbidities associated with CKD among YPLHIV. Our results call for more studies that characterise the spectrum and presentation of chronic comorbidities among YPLHIV with CKD for a more comprehensive understanding [5]. Comparison of YPLHIV with CKD with HIV negative controls might explain the contribution of HIV.

## Conclusion

The pattern of comorbidities among YPLHIV with CKD is uncertain and difficulties may relate to difficulty determining true kidney function and normal ranges in this population. Further studies are needed to discern the pattern of CKD complications to improve management efforts and clinical outcomes.

## Supporting information

Supplementary tables for cases with eGFR below 60ml

## Data Availability

All data produced in the present study are available upon reasonable request to the authors

## Ethical considerations

This study was approved by the Uganda Virus Research Institute (UVRI) Research Ethics Committee with reference number GC/127/946, the Uganda National Council of Science and Technology (reference number HS2578ES) and the London School of Hygiene and Tropical Medicine (reference 28797). Informed consent/assent was provided by all participants. Age-appropriate information about the study was provided prior to consent by the study nurses. All the participants aged more than 18 years provided a written informed consent. Those aged below 18 years (child) provided written assent and their caregivers provided a written informed consent. The studies were conducted in accordance with the declaration of Helsinki.

## Data availability

The data that support the findings of this study are available from the corresponding author upon reasonable request.

## Acknowledgments

We wish to acknowledge the participants, their caregivers and the facility staff where this study took place. We also acknowledge the study team, Miss Evelyn Natuha, Miss Stella Mirembe and Mr.

Malcolm Lwanga. We acknowledge the KCCA leadership and the PEPFAR program that supports HIV care for the YPLHIV.

## Supplementary material

There are two tables in the supplementary material section that show the sub analysis of cases that had an eGFR <60ml/min/1.73m2.

Supplementary table 1: Demographic and clinical characteristics of the cases with eGFR less than 60ml/min/1.73m^2^

Supplementary table 2: Distribution of comorbidities in the cases with eGFR less than 60ml/min/1.73m2

