## Supplementary tables for cases with eGFR below 60ml for "Comorbidities associated with chronic kidney disease among young people living with HIV in Uganda. A nested case control study"

Supplementary table 1 Demographic and clinical characteristics of the cases with eGFR less than 60ml/min/1.73m^2^

| **Variable** | **Total N=7** |
| --- | --- |
| **Sex** |  |
| Male | 5 (71.4) |
| Female | 2 (28.6) |
| **Age categorized** |  |
| Children | 5 (71.4) |
| Adults | 2 (28.6) |
| **Age in years (mean, SD)** | 16.6 (2.9) |
| **Social Economic Status** |  |
| Lowest | 3 (42.8) |
| Middle | 2 (28.6) |
| Highest | 2 (28.6) |
| **Is child in school** |  |
| No | 3 (42.8) |
| Yes | 4 (57.1) |
| **Body mass Index** |  |
| underweight | 3 (42.8) |
| Normal | 3 (42.8) |
| Overweight | 1 (14.2) |
| **Mid upper arm circumference** |  |
| Normal | 4 (57.1) |
| Malnourished | 3 (42.9) |
| **Weight in Kg Mean (SD)** | 51.9 (19.3) |
| **Muscle mass** |  |
| Normal muscle mass | 6 (85.7) |
| Abnormal muscle mass | 1 (14.3) |
| **TDF based regimen** |  |
| No TDF or ATV | 2 (28.6) |
| On TDF or ATV | 5 (71.4) |
| **Virally suppressed (VL <1,000 copies per ml)** |  |
| Yes | 6 (85.7) |
| No | 1 (14.3) |
| **Proteinuria** |  |
| Negative | 4 (57.1) |
| Positive | 3 (42.9) |
| **Albumin creatinine ratio** |  |
| Below 30mg/g | 2 (28.6) |
| Above 30mg/g | 5 (71.4) |

**Supplementary table 2 Distribution of comorbidities in the cases with eGFR less than 60ml/min/1.73m^2^**

| **Variable** | **Total**  **n=292** |
| --- | --- |
| **Blood pressure** |  |
| Systolic blood pressure ^a^ | 107.7 (11.1) |
| Diastolic blood pressure ^a^ | 70.4 (14.2) |
| Normal | 6 (85.7) |
| Hypertensive | 1 (14.3) |
| **Blood Sugar** |  |
| Blood glucose ^a^ | 88.4 (15.2) |
| Normal | 5 (71.4) |
| Prediabetic | 2 (28.6) |
| **Anaemia status** |  |
| Haemoglobin ^a^ | 13.1 (0.6) |
| No anaemia | 6 (85.7) |
| Anaemic | 1 (14.3) |
| **Serum Ferritin** |  |
| Ferritin ^b^ | 76.4 (113.1) |
| Normal | 6 (85.7) |
| High ferritin | 1 (14.3) |
| **Red blood cells** |  |
| Absolute count | 4.6 (0.4) |
| Normal count | 4 (57.1) |
| Low count | 3 (42.9) |
| **Haematocrit** |  |
| Haematocrit ^a^ | 40.5 (3.0) |
| Normal | 7 (100) |
| **Mean Corpuscular volume (MCV)** | |
| MCV ^a^ | 88.8 (6.7) |
| Normal | 6 (85.7) |
| Low MCV | 1 (14.3) |
| **Mean Corpuscular haemoglobin (MCH)** | |
| MCH ^a^ | 28.9 (2.2) |
| Normal MCH | 7 (100) |
| **White blood cell count** |  |
| Absolute ^b^ | 4.3 (1.3) |
| Normal | 5 (71.4) |
| Leukopenia | 2 (28.6) |
| **Platelets** |  |
| Count ^a^ | 333.1 (76.0) |
| Normal | 6 (85.7) |
| Thrombocytosis | 1 (14.3) |
| **Parathyroid hormone** |  |
| PTH ^b^ | 67.0 (18.3) |
| Normal parathyroid | 3 (42.8) |
| Hyperparathyroidism | 4 (57.1) |
| **Serum Calcium** |  |
| Calcium ^a^ | 2.3 (0.04) |
| Normal | 6 (85.7) |
| Hypocalcaemia | 1 (14.3) |
| **Serum Phosphate** |  |
| Phosphate ^b^ | 1.3 (0.3) |
| Normal | 6 (85.7) |
| Hyperphosphatemia | 1 (14.3) |
| **Blood Urea Nitrogen** |  |
| BUN ^a^ | 2.5 (0.7) |
| Normal | 5 (71.4) |
| Hypouremia | 2 (28.6) |
| **Serum Potassium** |  |
| Potassium ^a^ | 4.7 (0.4) |
| Normal | 7 (100) |
| **Serum Chloride** |  |
| Chloride ^a^ | 105.6 (3.3) |
| Normal | 6 (85.7) |
| Hyperchloremia | 1 (14.3) |
| **Serum Bicarbonate** |  |
| Bicarbonate ^a^ | 18.1 (1.7) |
| Low bicarbonate | 7 (100) |
| **Stunting** |  |
| Not stunted | 4 (80.0) |
| Stunted | 1 (20.0) |
| **Cognition** |  |
| Normal | 2 (28.6) |
| Impaired | 5 (71.4) |
